# High prevalence of SARS-CoV-2 antibodies in care homes affected by COVID-19; prospective cohort study, England

**DOI:** 10.1101/2020.08.10.20171413

**Authors:** Shamez N Ladhani, Anna Jeffery-Smith, Monika Patel, Roshni Janarthanan, Jonathan Fok, Emma Crawley-Boevey, Amoolya Vusirikala, Elena Fernandez, Marina Sanchez Perez, Suzanne Tang, Kate Dun-Campbell, Edward Wynne-Evans, Anita Bell, Bharat Patel, Zahin Amin-Chowdhury, Felicity Aiano, Karthik Paranthaman, Thomas Ma, Maria Saavedra-Campos, Joanna Ellis, Meera Chand, Kevin Brown, Mary E Ramsay, Susan Hopkins, Nandini Shetty, J. Yimmy Chow, Robin Gopal, Maria Zambon

## Abstract

**Background:** We investigated six London care homes experiencing a COVID-19 outbreak and found very high rates of SARS-CoV-2 infection among residents and staff. Here we report follow-up serological analysis in these care homes five weeks later.

**Methods:** Residents and staff had a convalescent blood sample for SARS-CoV-2 antibody levels and neutralising antibodies by SARS-COV-2 RT-PCR five weeks after the primary COVID-19 outbreak investigation.

**Results:** Of the 518 residents and staff in the initial investigation, 208/241 (86.3%) surviving residents and 186/254 (73.2%) staff underwent serological testing. Almost all SARS-CoV-2 RT-PCR positive residents and staff were antibody positive five weeks later, whether symptomatic (residents 35/35, 100%; staff, 22/22, 100%) or asymptomatic (residents 32/33, 97.0%; staff 21/22, 95.1%). Symptomatic but SARS-CoV-2 RT-PCR negative residents and staff also had high seropositivity rates (residents 23/27, 85.2%; staff 18/21, 85.7%), as did asymptomatic RT-PCR negative individuals (residents 62/92, 67.3%; staff 95/143, 66.4%). Neutralising antibody was present in 118/132 (89.4%) seropositive individuals and was not associated with age or symptoms. Ten residents (10/108, 9.3%) remained RT-PCR positive, but with lower RT-PCR cycle threshold values; all 7 tested were seropositive. New infections were detected in three residents and one staff member.

**Conclusions:** RT-PCR testing for SARS-CoV-2 significantly underestimates the true extent of an outbreak in institutional settings. Elderly frail residents and younger healthier staff were equally able to mount robust and neutralizing antibody responses to SARS-CoV-2. More than two-thirds of residents and staff members had detectable antibodies against SARS-CoV-2 irrespective of their nasal swab RT-PCR positivity or symptoms status.

## Introduction

Nursing and residential homes have been disproportionally affected by COVID-19 with high rates of hospitalisations and deaths among residents because of their advanced age and underlying comorbidities.^1,2^ In England, the first cases of imported COVID-19 cases were confirmed in late January 2020 with autochthonous transmission established by early March 2020. Cases peaked in mid-April before declining as a consequence of intense control measures.^3^ London, England, was one of the most affected cities and large outbreaks associated with high case fatality rates (CFR) among residents were reported in London care homes.^3^

During 10-13 April 2020, we investigated six London care homes reporting a suspected or confirmed COVID-19 outbreak to Public Health England (PHE).^4^ We found that 105 (40%) of 264 residents and 52 (21%) of staff had confirmed SARS-CoV-2, with half of both groups remaining asymptomatic throughout the surveillance period.^4^ Mass serological testing can help uncover the true extent of an outbreak within the care home setting,^5^ and potentially inform staff allocation and cohorting practices. Neutralising antibodies, in addition, may provide evidence for protection against reinfection, especially among the older residents who may not reliably mount an adequate protective response despite antibody production because of immunosenescence.^6,7^ As part of follow-up investigations, the residents and staff in the six care homes experiencing a COVID-19 outbreak were followed-up with a repeat nasal swab and a blood sample five weeks after the initial investigations. The study aimed to estimate SARS-CoV-2 seropositivity and neutralising antibodies in care home residents and staff of care homes experiencing a COVID-19 outbreak and assess any association with age, symptoms and SARS-CoV-2 RT-PCR positivity.

## Methods

The enhanced outbreak investigation initiated in April 2020 included six nursing and residential homes of different sizes, providing care for 43-100 residents with 14-130 staff per care home, with a confirmed COVID-19 outbreak.^4^ During the initial investigation, nasal swabs were taken for SARS-CoV-2 RT-PCR for all residents and staff working in the care home at the time. Infection control measures were reinforced and all SARS-CoV-2 RT-PCR positive individuals were isolated. All tested participants were followed up for any symptoms during the two weeks before, at the time of testing and for two weeks after the test.^4^

Follow-up investigation involved a repeat nasal swab and a blood sample from all participants five weeks after the initial RT-PCR testing. Consent was obtained by care home managers from staff members and residents or their next of kin as appropriate. Testing began on the week of May 18, 2020. Care home staff took nasal swabs for residents and submitted their own samples by self-swabbing with appropriate instructions. Care home nurses took blood samples from residents and their colleagues, with external phlebotomists assisting two care homes with sampling. It was not appropriate or possible to involve patients or the public in the design, or conduct, or reporting, or dissemination plans of our research

### SARS-CoV-2 antibody testing

SARS-CoV-2 Infected virus lysate assay: Native virus antigen ELISA was modified from a previously described MERS-CoV assay.^8^ Microplate bound detergent extracted lysates of SARS-CoV-2 (isolate England/02/2020) infected Vero E6 cells and uninfected cells were reacted with a serial dilution of convalescent serum obtained from participants in an indirect ELISA format. Virus lysates contained a mixture of viral proteins expressed in Vero E6 cells, including viral nucleocapsid and spike proteins.

### Microneutralisation assay and neutralising antibody titre

SARS-CoV-2 (isolate England/02/2020) specific neutralising antibody levels were measured using a modification of the WHO influenza microneutralisation methodology.^9^ Briefly, 200 TCID 50 of virus was incubated with serial dilutions of serum from participants, after which a suspension of Vero E6 cells were added. After 22 hours, cells were fixed and in-cell SARS-CoV-2 nucleoprotein (NP) expression determined by ELISA. The virus neutralising antibody titre was determined as the serum concentration that that inhibited 50% SARS-CoV-2 NP expression. All work was undertaken in a BSL-3 laboratory.

### SARS-CoV-2 PCR

Nucleic acid was extracted from samples and analysed by a real-time reverse transcription (RT) PCR assay targeting a conserved region of the open reading frame (ORF1ab) gene of SARS CoV-2, together with detection of an assay internal control to monitor the extraction and RT-PCR processes. This assay used the primers and probe sequences made public by CDC China (http://ivdc.chinacdc.cn/kviz/202001/t20200121_211337.html) and required 5μL RNA in a total RT-PCR reaction volume of 25μL. Reverse transcription and PCR amplification was performed on an Applied Biosystems 7500 FAST system.

### Statistical analysis

Descriptive analyses were performed. Data that did not follow a normal distribution were described as medians with interquartile ranges and compared using the Mann Whitney U test. Antibody concentrations were presented as ELISA index values with medians and 95% confidence intervals (95% CI). Antibody concentrations above the index value of 0.5 were considered positive. Median antibody concentrations were compared using Kruskal-Wallis with Dunn’s multiple comparisons test adjustment. Categorical variables were described as proportions and compared using chi squared or Fishers Exact test as appropriate. Data were analysed using Stata version 15.0 (Statcorp, Tx) and GraphPad Prism.

## RESULTS

### Seropositivity

Of the original 518 residents and staff involved in the initial care home outbreak investigation during 10-13 April 2020, 394 (76.1%) consented for follow-up investigations at median of 36 days (range, 30-45 days) (**Figure 1**). SARS-CoV-2 seropositivity for the cohort was 77.9% (95%CI, 73.6-81.7%; **Table 1, Figure 2a**). Seropositivity was associated with being symptomatic and SARS-CoV-2 RT-PCR nasal swab positive during at the initial test, but not with gender, age or being a resident or staff member (**Table 1, Figure 2**).

**Figure 1.**
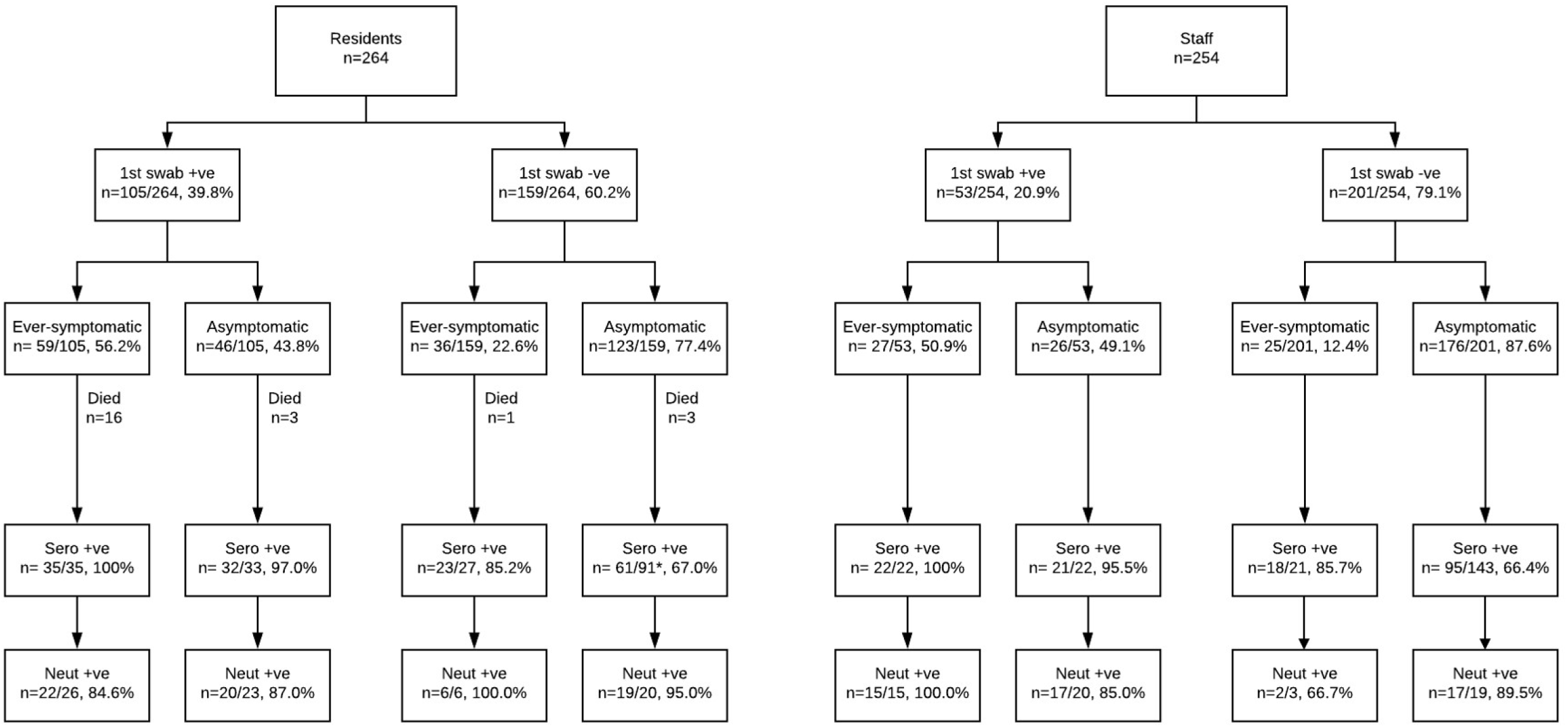
Flow diagram of residents and staff in 6 London care homes experiencing a COVID-19 outbreak during the pandemic who consented to follow-up testing including blood sampling for SARS-CoV-2 antibodies four to six weeks later. ‘Ever-symptomatic’ indicates that symptoms were experienced at some point during the follow-up period. *Four individuals in this group became SARS-CoV-2 PCR positive at follow-up RT-PCR testing conducted simultaneously with SARS-CoV-2 antibody testing.

**Table 1:** Demographics of care homes cohort and seropositivity by group: percentage and 95% confidence intervals shown. Statistical analysis using ^1^Fisher’s exact test and ^2^Chi square test of proportions, p values as shown.

|  | n | % total | Seropositive (n) | (%) | 95% CI |
| --- | --- | --- | --- | --- | --- |
| Overall | 394 | 100 | 307 | 77.9 | 73.6 - 81.7 |
| Sex |  |  |  |  |  |
| Male | 95 | 24.1 | 73 | 76.8 | 67.4 - 84.2 |
| Female | 299 | 75.9 | 234 | 78.3 | 73.2 - 82.6 |
| p value |  |  |  | 0.78<sup>1</sup> |  |
| Age group |  |  |  |  |  |
| 19-39 | 62 | 15.7 | 46 | 74.2 | 62.1 – 83.5 |
| 40-59 | 117 | 29.7 | 91 | 77.8 | 69.4 – 84.4 |
| 60-79 | 68 | 17.3 | 49 | 72.1 | 60.4 – 81.3 |
| >80 | 147 | 37.3 | 121 | 82.3 | 75.4 – 87.6 |
| p value |  |  |  | 0.31<sup>2</sup> |  |
| Residents and Staff |  |  |  |  |  |
| Residents | 186 | 47.2 | 151 | 81.2 | 75.0 - 86.2 |
| Staff | 208 | 52.8 | 156 | 75.0 | 68.7 - 80.4 |
| p value |  |  |  | 0.15<sup>1</sup> |  |
| Symptoms at any time during first test period |  |  |  |  |  |
| Symptomatic | 105 | 26.6 | 98 | 93.3 | 86.9 - 96.7 |
| Asymptomatic | 289 | 73.4 | 209 | 72.3 | 66.9 - 77.2 |
| p value |  |  |  | <0.0001<sup>1</sup> |  |
| First round PCR results |  |  |  |  |  |
| 1st PCR + | 112 | 28.4 | 110 | 98.2 | 93.7 - 99.7 |
| 1st PCR - | 285 | 72.3 | 197 | 69.1 | 63.5 - 74.2 |
| p value |  |  |  | <0.0001<sup>1</sup> |  |

**Figure 2.**
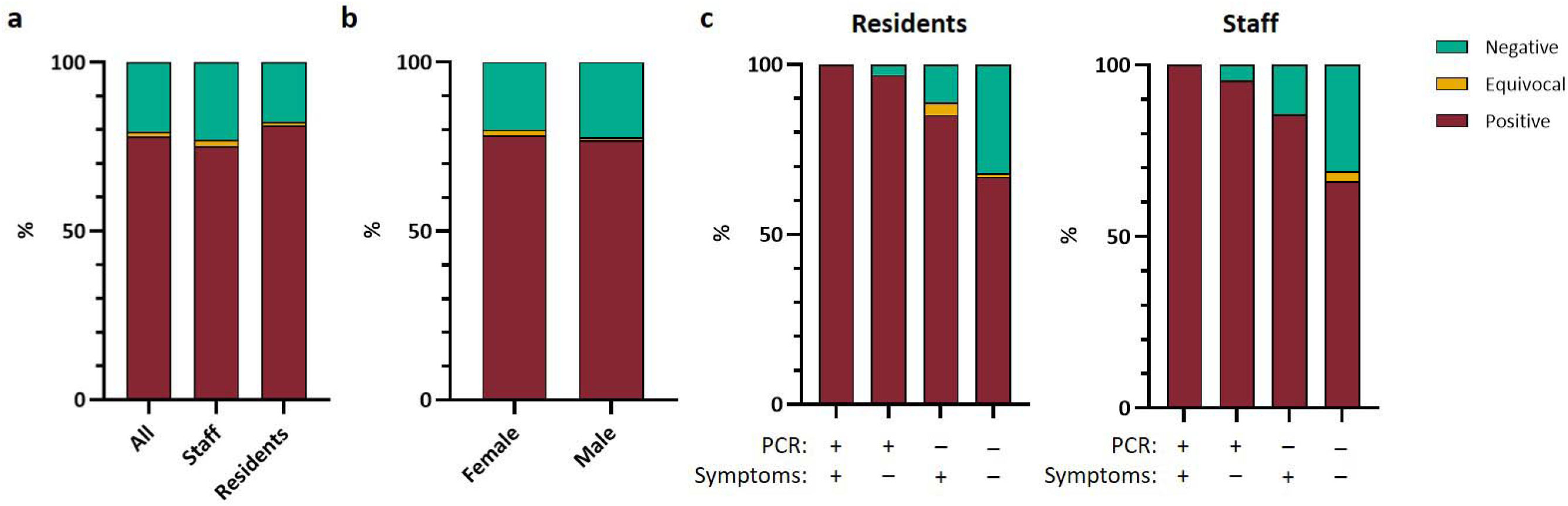
Proportion of indicated study populations with positive (maroon), equivocal (yellow) or negative (green) native viral antigen lysate assay, a) Seroconversion for all cohort and staff and resident sub groups, b) Seroconversion by sex for whole cohort, c) Seroconversion b y acute RT-PCR result and presence of symptoms for residents and staff. All N = 394; Staff N= 208; Residents N= 186.

### Residents

Twenty-one of the 264 residents tested in the initial investigations died within two weeks and two others died prior to follow-up testing. Thus, 186 of the remaining 241 residents who consented to SARS-CoV-2 antibody testing and 81.2% (151/186) were seropositive. Of the 35 residents who had been symptomatic during the outbreak and were PCR-positive during the initial testing period, all were SARS-CoV-2 antibody positive (**Figure 1, Figure 2c**). At the same time, 97% (32/33) of residents who were PCR-positive but remained asymptomatic throughout the outbreak were also SARS-CoV-2 antibody positive (**Figure 1, Figure 2c**). Of the 118 residents who had tested PCR-negative initially, 85.2% (23/27) of those who had been symptomatic during the outbreak were SARS-CoV-2 antibody positive and 67.0% (61/91) of residents who remained asymptomatic (**Figure 1, Figure 2c**).

### Staff

Among the 254 staff members involved in the initial investigation, 208 consented to additional investigations and 75.0% (156/208) were seropositive (**Figure 2a**). All 22 staff members who were SARS-CoV-2 RT-PCR positive at initial testing and symptomatic during the outbreak were positive for SARS-CoV-2 antibodies, as were 95.5% (21/22) of RT-PCR positive asymptomatic staff (**Figure 1, Figure 2c**). There were 201 staff members who were SARS-CoV-2 RT-PCR negative and 25 went on to develop COVID-19 compatible symptoms during the follow-up period; of the21 staff tested, 18 (85.7%) were SARS-CoV-2 antibody positive. The remaining 176 SARS-CoV-2 PCR negative staff members were asymptomatic throughout the surveillance period and, of the 143 tested, 95 (66.4%) were SARS-CoV-2 antibody positive (**Figure 1, Figure 2c**).

### SARS-CoV-2 Antibody seropositivity and index values

There was no association between SARS-CoV-2 seropositivity and age (Chi-square test, P=0.31) (**Figure 3, left panel**). Among SARS-CoV-2 antibody positive individuals, there was no significant difference in median index value between the those who had been PCR positive or negative at initial testing (**Figure 3, right panel**). There was no association between median SARS-CoV-2 antibody index value and symptom status, PCR status or gender (data not shown). Six individuals had equivocal SARS-CoV-2 antibody index values; all were asymptomatic throughout the surveillance period and were SARS-CoV-2 RT-PCR negative on nasal swabs at both timepoints.

**Figure 3.**
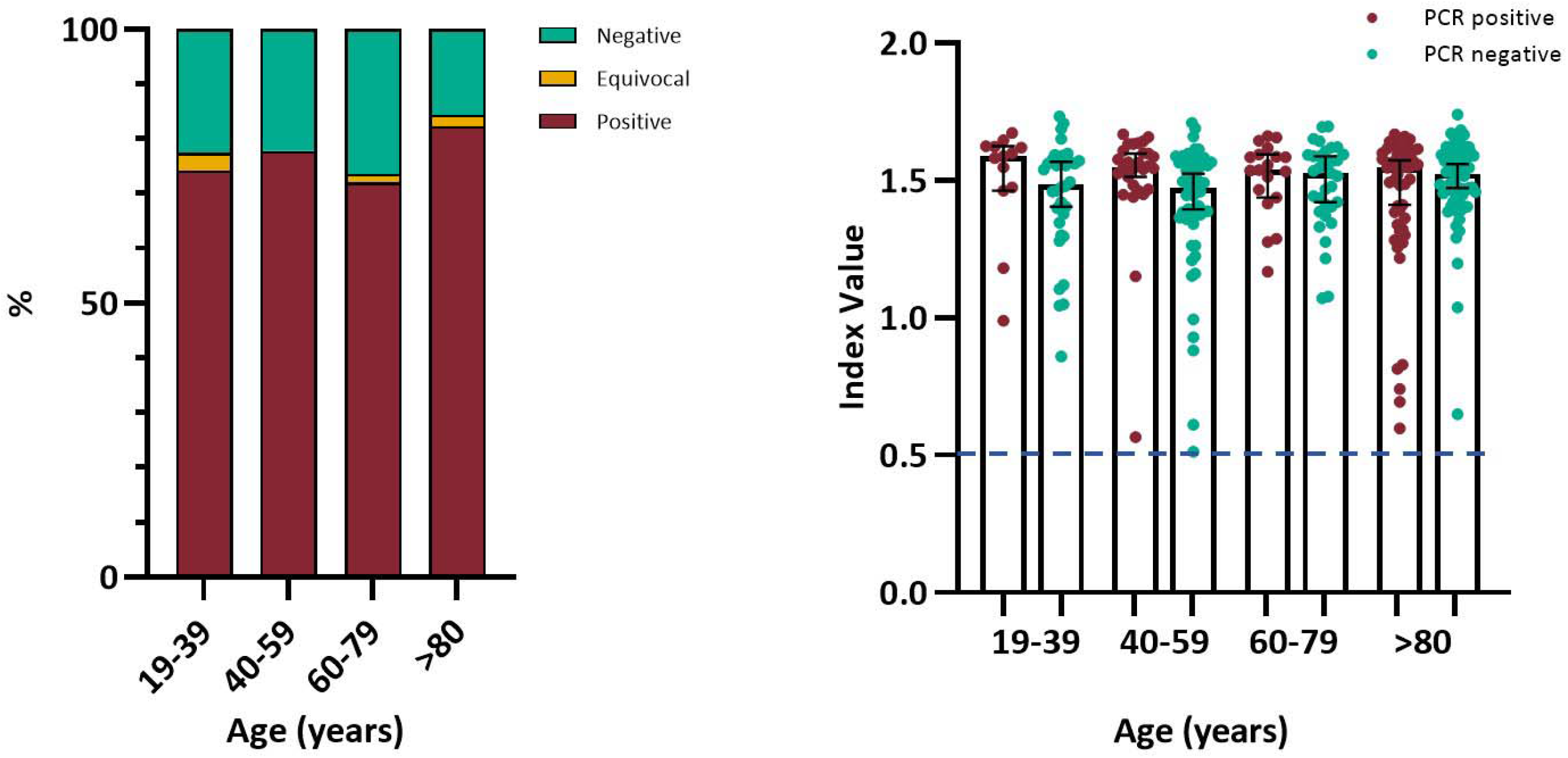
Age group analysis native viral antigen lysate assay. Left panel: Seroconversion percentage by age group for whole cohort, N=394. Right panel: Native viral antigen assay index value for seropositive individuals by initial RT-PCR status and age group, N=307. Bars indicate median and 95% confidence interval. Dashed line indicates assay positive cut-off. Statistical analysis using Chi square test of proportion and Kruskal-Wa Mis with Dunn’s multiple comparisons test, no significant difference.

### Neutralising antibodies

Neutralising antibodies were detected in 89.4% (118/132) of seropositive individuals. There was no association between the detection of SARS-CoV-2 neutralising antibodies and age, sex, symptom status or PCR status (**Figures 4a-c**, data not shown). There was a trend toward increasing neutralising antibody titre with age (**Figure 4d**).

**Figure 4.**
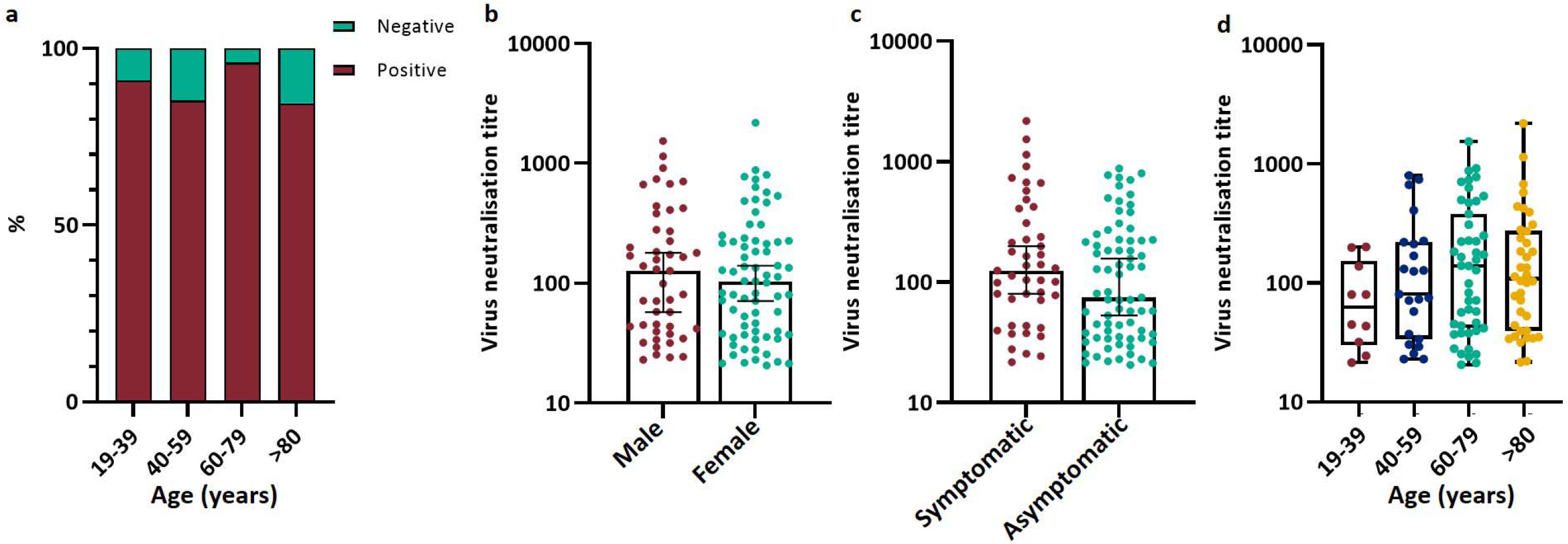
Virus neutralising antibody titre analysis, a) Virus neutralising positive and negative percentage by age group for whole cohort. N=132 b) Virus neutralisation titre by sex. Bars indicate median and 95% confidence interval, c) Virus neutralisation titre by symptom status during the initial testing period. Bars indicate median and 95% confidence interval, d) Virus neutralisation titre by age group. Box and whisker plot with bars indicating full range of results. N=118. Statistical analysis a) chi-square test, no significant difference; b and c) Mann-Whitney U Test; d) Kruskal Wa Mis with Dunn’s multiple comparisons test adjustment, no significant difference.

### Nasal Swab RT-PCR

All consenting residents and staff had a repeat nasal swab at the time of blood sampling. Thirteen residents were SARS-CoV-2 RT-PCR positive, including 10 who had been SARS-CoV-2 RT-PCR positive at an interval of 36-45 days previously, although SARS-CoV-2 RT-PCR Ct values were significantly lower at follow-up (**Figure 5**). Of these ten, 7 also had serum samples taken and all were seropositive (**Table 2**). Four residents became SARS-CoV-2 RT-PCR positive at follow-up and were seropositive (**Figure 1**); all remained asymptomatic. None of the SARS-CoV-2 RT-PCR positive staff at the initial visit were positive on repeat testing. A previously SARS-CoV-2 RT-PCR negative staff member who was asymptomatic throughout the surveillance period became positive on repeat testing, with a Ct value of 35.6; this staff member was also seropositive for SARS-CoV-2 antibodies.

## DISCUSSION

In six London care homes experiencing a COVID-19 outbreak at the peak of the pandemic, 81.2% of surviving residents and 75.0% of staff were SARS-CoV-2 antibody positive five weeks after the initial outbreak investigation. These rates are far higher than any other cohort including frontline healthcare workers managing patients with confirmed COVID-19 in hospitals.^10-12^ Almost all residents and staff who were SARS-CoV-2 RT-PCR positive on nasal swab at initial testing developed SARS-CoV-2 antibodies, irrespective of whether they were symptomatic at any point during the outbreak. High seropositivity rates were also observed for symptomatic residents and staff even if they had a previously negative SARS-CoV-2 RT-PCR test. The serological investigation emphasises the extent to which SARS CoV-2 can penetrate vulnerable communities in closed settings, and the underestimation of exposure through point prevalence estimates using RT-PCR from nasal swabs. The lowest seroprevalence was observed in residents and staff who remained asymptomatic throughout the outbreak and who were SARS-CoV-2 RT-PCR negative on both testing visits but, even in this cohort, more than two-thirds were positive for SARS-CoV-2 antibodies. In this cohort, SARS-CoV-2 antibody levels were not associated with age, sex, presence of symptoms, PCR-positivity or resident/staff status.

There are now sensitive and specific serological assays, such as the one used in this investigation based on using native viral antigens derived from infected cells.^13^ Overall, a recent systematic review estimated that serological tests had 30% sensitivity for SARS-CoV-2 antibodies during the first week after symptom onset, rising to 72% in the second week, 91% in the third week and 96% up to 5 weeks later.^14^ The finding that almost all residents and staff with confirmed COVID-19 through nasal swab SARS-CoV-2 RT-PCR – irrespective of whether they were ever experienced symptoms during the outbreak – is reassuring and validates the use of our serological assay as a measure of past exposure. The very high seropositivity rates among symptomatic but RT-PCR negative residents and staff suggests that their original illness was also most likely due to COVID-19 and highlights the sensitivity limitations of single point nasal swabbing for diagnosis and the narrow window of SARS-CoV-2 detection in infected individuals, and significantly underestimated exposure to SARS-CoV-2 in these outbreak settings.^15^ The very high seropositivity of 75.0% among care home staff compared to 17-44% of patient-facing healthcare workers is staggering.^10,11^ A possible explanation may be more prolonged and intense exposure to the virus because of level of care required by the residents.^2,15^ In our initial investigations, we also found evidence of transmission between staff members in care homes, highlighting the importance of maintaining infection control practices for all contact, including those between staff, whilst on care home premises.^8^ Despite reinforcement of infection control measures at the outset, one further staff member and three residents became infected with SARS-CoV-2 at follow-up. Residents and staff who were SARS-CoV-2 RT-PCR positive at follow-up all had high Ct values, consistent with non-viable virus at the time of testing,^4^ and were also SARS-CoV-2 antibody positive. Further studies are needed to assess whether the presence of SARS-CoV-2 antibodies, including neutralising antibodies, are protective against re-infection and, if so, the duration of protection. Questions also remain as to whether antibody concentrations provide a useful measure of protection in infected individuals.^16^

Notable, too in this study, was the observation that SARS-CoV-2 antibody levels were similar among symptomatic and asymptomatic residents and staff across the care homes, which contrasts with recent reports suggesting that higher antibody levels and persistence were achieved in patients with more severe disease compared to those with mild disease or asymptomatic infection.^12^ The high fatality rates among residents across the six care homes, particularly affecting those who had been symptomatic and SARS CoV-2 RT-PCR positive indicates that that the cohort described here is more representative of milder illness and depleted of individuals who suffered the most severe outcomes of infection.^4^ We also found that 90% of seropositive staff and residents had neutralising antibody responses, with no significant differences in neutralising antibody levels between by age, sex, symptom status or staff/resident status. There was a trend towards increasing neutralising antibody titres with increasing age (Figure 4c) but this was not statistically significant. These findings of such robust antibody responses in surviving care home residents, especially when compared to younger, healthier staff members with similar exposure risks to SARS-CoV-2, are novel and may play an important part in future recommendations for infection control practices and vaccination against SARS-CoV-2.

Several key questions relating to this novel pandemic remain to be answered and are particularly relevant to this highly vulnerable population and setting. In particular, it is not known whether SARS-CoV-2 antibodies are protective against re-infection.^16^ We identified a small number of residents who were still RT-PCR positive up to five weeks later; all had detectable antibodies, including some with neutralising antibodies at the time of the persistent virus detection.^17^ The RT-PCR Ct values were consistent with non-live virus in all residents and staff members who were RT-PCR positive on nasal swab at follow-up. The prolonged nasal swab RT-PCR positivity in a proportion of residents and staff is consistent with a recent large healthcare worker study where up to a quarter were still RT-PCR positive up to six weeks later, highlighting yet another limitation of our understanding of the kinetics of viral replication and immune responses COVID-19.^18,19^

The strengths of our investigations include the extensive and robust epidemiological, virological and serological testing of residents and staff across six London care homes experiencing large outbreaks of COVID-19, the broad age ranges involved, the daily follow-up after initial testing and the high uptake for retesting five weeks later. The data collected have provided a wealth of information on SARS-CoV-2 infection, transmission and antibody responses in a high-risk care setting involving a very vulnerable cohort. A limitation is that the care homes were already in the middle of the outbreak. Consequently, some residents had already developed COVID-19 and some had died of their infection. Another limitation was that we did not obtain blood samples for antibody testing at the first visit, which could have provided additional useful information on antibody kinetics in a large cohort of elderly residents and younger staff members. The lower nasal swab positivity during the initial investigations compared to the antibody results five weeks later reflects the limited sensitivity of test, the quality of sampling, the stage of infection at the time of testing and the gene targets used by different RT-PCR assays.^20^ Some of these limitations could potentially have been mitigated by repeated swabbing at different time points.

In conclusion, almost all residents and staff with confirmed SARS-CoV-2 infection had detectable antibodies five weeks later, irrespective of whether they were ever symptomatic or remained asymptomatic throughout the outbreak. Additionally, a high proportion of those who were symptomatic but SARS-CoV-2 RT-PCR negative were also seropositive. SARS-CoV-2 antibody levels were not associated with age, sex, PCR positivity, symptomatic/asymptomatic or resident/staff status. Our findings demonstrate the older and vulnerable residents are able to mount a robust antibody response to SARS-CoV-2 that is similar to younger and healthier staff members. Further studies are needed to determine whether SARS-CoV-2 antibodies protect against re-infection and, if so, the duration of protection.

## Data Availability

Additional data are available for sharing

## Ethics approval

The research protocol was approved by the PHE Research Ethics and Governance Group (REGG Ref: NR0204, 07 May 2020).

## Acknowledgements

The authors are very grateful to the care home managers, their staff and the residents for their willingness to support the investigation, along with the staff in the immunisation and countermeasures division, PHE Operations, the virus reference department, the London Coronavirus Response Cell and Field Services for their help and support with the investigation.

## Author Contribution

SNL, JYC and MZ conceived and designed the study; all authors contributed to the analysis and interpretation of the data, and commented on the manuscript prior to submission

## License statement

*“The Corresponding Author has the right to grant on behalf of all authors and does grant on behalf of all authors, a worldwide licence to the Publishers and its licensees in perpetuity, in all forms, formats and media (whether known now or created in the future), to i) publish, reproduce, distribute, display and store the Contribution, ii) translate the Contribution into other languages, create adaptations, reprints, include within collections and create summaries, extracts and/or, abstracts of the Contribution, Hi) create any other derivative work(s) based on the Contribution, iv) to exploit all subsidiary rights in the Contribution, v) the inclusion of electronic links from the Contribution to third party material where-ever it may be located; and, vi) licence any third party to do any or all of the above*.*”*

## Dissemination declaration

we have provided the overall findings of the study to the participating care homes

## Data Sharing

there are additional data for sharing

